# SWI and T2*-GRE Microhemorrhage Counts in Anti-Amyloid Therapy Eligibility: A Real-World-Calibrated Simulation Study

**DOI:** 10.64898/2026.06.22.26356283

**Authors:** Kenichiro Sato, Ryoko Ihara, Masanori Kurihara, Fumio Suzuki, Aya M. Tokumaru, Yoshiki Niimi, Takeshi Iwatsubo, Atsushi Iwata, Alzheimer’s Disease Neuroimaging Initiative

## Abstract

**INTRODUCTION:** Anti-amyloid therapy eligibility excludes patients with five or more cerebral microhemorrhages (CMHs), but current guidance allows either T2*-GRE or the more sensitive SWI. This may create sequence-dependent differences in eligibility classification.

**METHODS:** We fitted a Bayesian right-censored zero-inflated Poisson model to single-center real-world SWI-based CMH counts from 130 memory clinic patients. We then simulated T2*-GRE counts under a directional binomial detection model across a range of relative detection probabilities and estimated two metrics: *P(T2* <5 | SWI* ≥*5)* and *P(SWI* ≥*5 | T2* <5)*.

**RESULTS:** The model estimated a zero-inflation probability of 0.57 and a Poisson mean of 2.29 in the susceptible subpopulation. In the illustrative 60% relative-detection scenario, the model estimated nearly 80% under-detection among SWI-ineligible patients, whereas approximately 5% of T2*-GRE-eligible patients would have been classified as ineligible by SWI.

**DISCUSSION:** A uniform numeric CMH threshold may not be operationally equivalent across SWI and T2*-GRE. Standardized paired-sequence outcome studies are needed to determine whether SWI-defined and T2*-GRE-defined thresholds carry equivalent ARIA risk.

## Introduction

The recent approval of anti-amyloid antibodies (AATs), such as lecanemab and donanemab, marks a significant advancement in the treatment of early-stage Alzheimer’s disease (AD) [vanDyck2023; Sims2023]. While these therapies offer the first opportunity to modify the disease’s course, they are associated with safety concerns, most notably Amyloid-Related Imaging Abnormalities (ARIA) [Cogswell2022; Hampel2023; Zimmer2025]. A key criterion for minimizing these risks involves baseline magnetic resonance imaging (MRI) to screen for contraindications. Appropriate use recommendations [Cummings2023; Rabinovici2025] and Japan’s Optimal Use Guidelines (OUGs) [MHLW2023; MHLW2024; Iwata2024] for lecanemab and donanemab require that, patients with five or more cerebral microhemorrhages (CMHs) are to be excluded from treatment. This is a critical step in the patient selection process, because greater baseline hemorrhagic burden is associated with higher ARIA-H risk [Cogswell2022; Hampel2023; Zimmer2025]. A recent Alzheimer’s Association ARIA workgroup review also identified baseline microhemorrhages and cortical superficial siderosis as established ARIA risk factors, while emphasizing that ARIA risk is influenced by multiple factors beyond CMH count alone [vanEtten2026].

Current guidelines permit the use of either T2*-weighted gradient-recalled echo (T2*-GRE) or susceptibility-weighted imaging (SWI) sequences for CMH detection [Kakeda2025]. It is well-established that SWI has a significantly higher sensitivity for detecting CMHs compared to T2*-GRE [Nandigam2009; Cheng2013; Shams2015; Haller2021; Cogswell2022; Shams2015]. A recent ARIA workgroup review similarly noted that susceptibility-weighted sequences provide higher sensitivity than GRE and that use of SWI rather than GRE may change management in approximately 4% to 5% of patients [vanEtten2026]. This disparity in detection performance creates a clinical ambiguity: applying a uniform exclusion threshold of ≥ 5 baseline CMHs would make eligibility depend on the MRI sequence used, despite the exclusion criteria determined based on T2*-based evidence [Sima2025]. A patient who would be counted as having ≥5 CMHs on SWI at one site might register <5 on T2*-GRE at another site and therefore not meet the exclusion criterion and proceed to treatment. In real-world paired readings, discordance in the opposite direction may also occur because of reader variability, artifacts, or lesion mimics, although the primary model in this study focused on the directional effect of lower T2*-GRE detection relative to SWI. Although T2*-GRE had been dominantly used in the clinical trials, it has been reported that SWI is increasingly used in routine clinical practice [Shams2015;Sima2025]. As of late 2024, according to an online survey to dementia specialist in Japan, approximately half of specialists reported the use of SWI for brain MRI screening [Sato2025], reflecting that SWI-capable equipment is not universally available. Consequently, issues arising from the SWI–T2* performance discrepancy are expected to become more pronounced going forward in the clinical practice of AATs.

Given the direct impact on patient safety and access to treatment, the discrepancy between these imaging techniques poses a substantial challenge for the real-world implementation of AATs. Although prior simulation work and a recent ARIA workgroup review have highlighted the potential impact of sequence choice [Sima2025; vanEtten2026], and conference-level paired-sequence data from TRAILBLAZER-ALZ 6 have begun to address this issue [Svaldi2025], peer-reviewed paired-sequence evidence remains limited. In particular, the magnitude of sequence-dependent eligibility discordance remains insufficiently characterized in real-world clinical practice, especially in implementation settings where SWI is increasingly used for eligibility screening. This study, therefore, aimed to model the potential frequency of conflicting eligibility decisions arising from the choice between SWI and T2*-GRE. Using a Monte Carlo simulation parameterized with real-world SWI-based CMH count data obtained from our memory clinic practice, we estimated the probability of misclassification associated with current MRI protocols. In practice, two legitimate perspectives coexist. A safety-first view emphasizes strict adherence to the ≥5 microhemorrhage threshold, regardless of whether CMHs are counted on T2*-GRE or SWI, to minimize ARIA risk. In contrast, an access-preserving view notes that the clinical trials used T2*-GRE; therefore, directly transplanting the same numeric cutoff to the more sensitive SWI may over-exclude patients who were considered eligible under trial-based criteria. Rather than relying on one of these positions, our study quantifies the magnitude of discordance that clinicians must navigate when different sequences are used.

## Methods

### Study design and analytic cohort

The present simulation study used a subset of patients from the anti-amyloid therapy screening pathway at our center, which has been described previously [Ihara2026]. Briefly, candidates entered the pathway either through self-referral with a specific interest in anti-amyloid therapy or through physician referral during routine dementia care. First-stage screening was performed at the memory or neurology clinic to assess clinical stage, cognitive severity, major exclusion criteria, and willingness to undergo regular intravenous treatment. Patients who passed this stage attended the specialized disease-modifying therapy clinic, where physicians reconfirmed eligibility and provided detailed information about anti-amyloid therapy. Patients who remained willing to proceed then underwent the remaining eligibility assessments, including CDR assessment, brain MRI, and amyloid confirmation by amyloid PET or CSF biomarkers.

In the previously reported full implementation cohort [Ihara2026], 312 patients attended the disease-modifying therapy clinic; 93 did not proceed after the informed-consent step, and 219 proceeded to the remaining eligibility assessments. Among these, 88 were subsequently excluded, including 44 because of negative amyloid-beta accumulation and 33 because of MRI contraindications or acute cerebral infarction, and 131 ultimately initiated anti-amyloid therapy.

For the present analysis, we focused on the MRI-evaluated subset of this clinical pathway. We included consecutive patients who underwent baseline brain MRI with SWI on a 3.0T scanner (Ingenia Elition 3.0T, Philips) as part of the clinical workup for anti-amyloid therapy eligibility assessment. These patients included both those who ultimately initiated anti-amyloid therapy and those who did not. During the study period, 138 unique patients underwent baseline MRI with SWI for this purpose. SWI-based cerebral microhemorrhage counts were unavailable in 8 patients, who were excluded from model fitting. The final analytic cohort therefore comprised 130 unique patients with available SWI-based cerebral microhemorrhage counts. This cohort was used to estimate the baseline microhemorrhage count distribution for the simulation model.

For our analysis, CMH counts were recorded as integers, and counts of five or more had been recorded as a right-censored category (“≥ 5”). After excluding cases with missing in CMH data, the final dataset for model fitting comprised 130 unique patients, among whom 94 (72.3%) eventually received AAT infusion.

### Modeling the Baseline Microhemorrhage Distribution

The distribution of CMH counts in our clinical cohort exhibited two key characteristics: a high proportion of patients with zero CMHs (61.5%,) and right-censoring at five or more CMHs (Figure 1). To accurately capture these features, we fitted a Bayesian right-censored, zero-inflated Poisson (ZIP) model to the observed SWI-based CMH counts. The ZIP model estimates two fundamental parameters that describe the CMH distribution: λ (lambda), the mean rate of CMHs in the sub-population of patients who are susceptible to having them (i.e., those not in the “true zero” group), and this reflects the underlying risk of CMH burden. And zi (ζ), the zero-inflation probability, representing the proportion of patients who are structurally immune to developing CMHs or have a count of exactly zero, distinct from those who might have zero by chance from the Poisson distribution.

**Figure 1.**
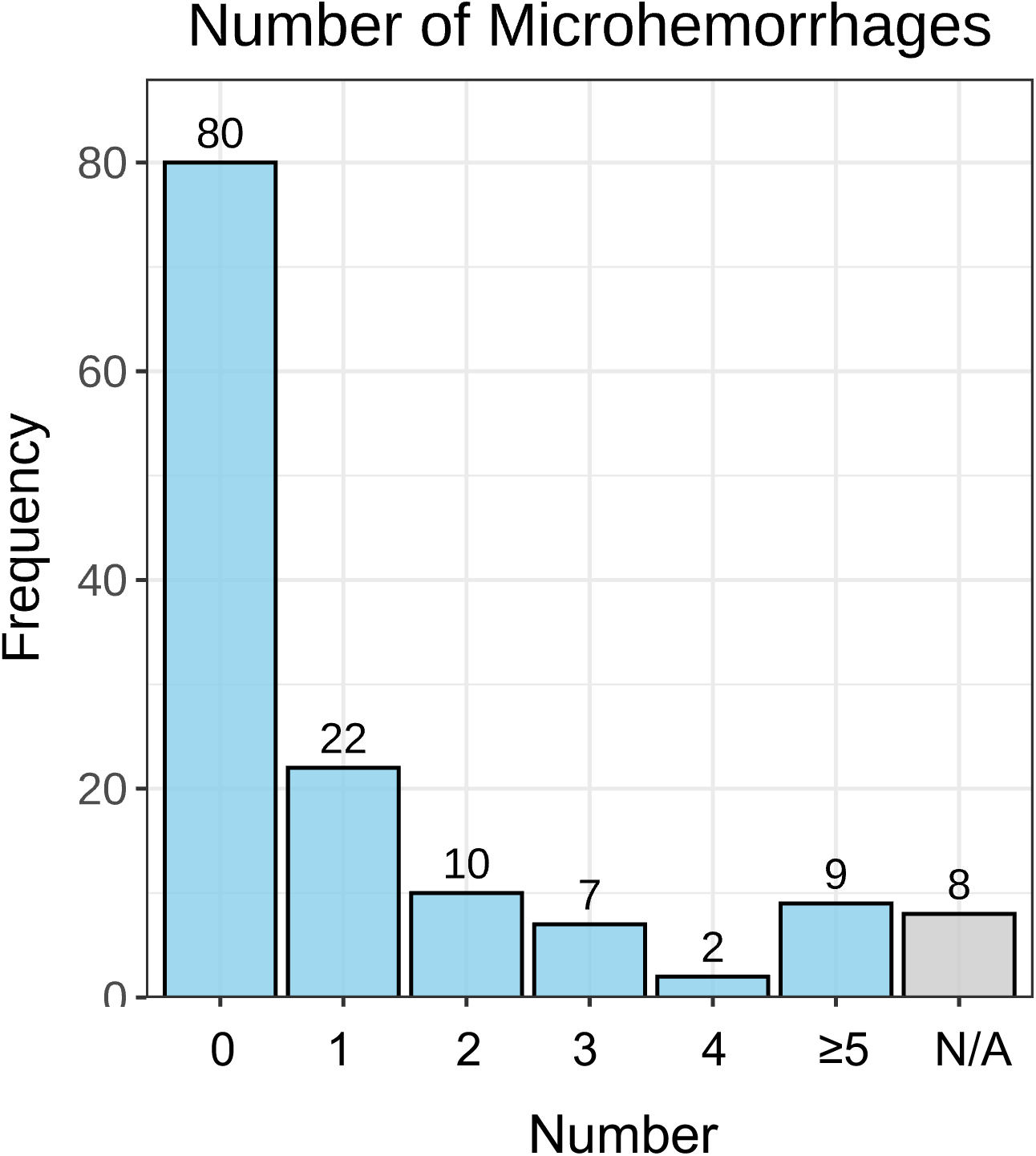
Distribution of SWI-based cerebral microhemorrhage counts in the clinical cohort. The histogram shows the distribution of cerebral microhemorrhage (CMH) counts determined from clinical readings of baseline SWI. The category labeled “≥5” represents five or more CMHs, because counts beyond the treatment-exclusion threshold were recorded as a right-censored category. The gray bar indicates cases with unavailable CMH count data; these cases were excluded from model fitting. After excluding these missing cases, 130 patients were included in the Bayesian right-censored zero-inflated Poisson model. **Abbreviations**: CMH, cerebral microhemorrhage; SWI, susceptibility-weighted imaging.

The model was implemented in the R software (version 4.3.1) using the package {*brms*} for Bayesian analysis. We used default, weakly informative priors and ran four parallel Markov Chain Monte Carlo (MCMC) chains for 4,000 iterations each, with a warm-up period of 1,000 iterations, to ensure convergence. Model convergence was confirmed by examining trace plots and ensuring the Gelman-Rubin diagnostic (G^⏢^) was equal to 1.00 for all parameters.

### Monte Carlo Simulation Framework

With the posterior distributions for λ and ζ established, we designed a Monte-Carlo simulation to evaluate the impact of using a less sensitive MRI sequence. The core assumption of our analysis is that SWI-based CMH counts represent the most accurate, or “true,” biological burden of microhemorrhages.

The simulation proceeded as follows. First, we generate “true” CMH counts: a large virtual cohort of 10,000 patients was generated. For each fixed value of *p_detect_*, we repeatedly generated virtual cohorts by drawing λ and zi from their posterior distributions. For each posterior draw, a “true” SWI-based CMH count (*Y_SWI_*) was simulated from the fitted ZIP distribution.

The simulated T2*-GRE count (*Y_T2*_*) was then generated by binomial sampling from *Y_SWI_*. Outcome measures were summarized as posterior medians and 95% credible intervals. The relative detection power of T2*-GRE, denoted as pdetect, represented the probability that any single CMH visible on SWI was also detected by T2*-GRE. Thus, for each virtual patient, the simulated T2*-GRE count was generated as *Y_T2*_ ∼ Binomial(n = Y_SWI_, p = p_detect_)*. We refer to this as the primary directional detection model, because T2*-GRE was modeled as detecting a subset of SWI-visible CMHs. This model isolates the effect of lower T2*-GRE detection but does not represent reverse discordance due to artifacts, lesion mimics, or reader variability.

We then varied the relative detection power of T2*-GRE. To assess a broad range of potential clinical scenarios, we varied pdetect from 0.30 to 1.00 in discrete steps. A value of 1.00 represents perfect agreement between the sequences, while lower values reflect poorer T2*-GRE sensitivity. Prior comparative MRI studies have shown higher CMH detection with SWI than with T2*-GRE, but the relative performance of T2*-GRE varies by slice thickness, field strength, acquisition parameters, and reading rules [Nandigam2009; Cheng2013; Shams2015]. We therefore interpreted the full range of pdetect values rather than assuming a single true value. The *p_detect_* = 0.60 scenario was used only as an illustrative example in the text. For each pdetect level, every virtual patient was categorized based on whether their *Y_SWI_* and *Y_T2*_* counts were below or above the exclusion threshold of five CMHs.

### Outcome Measures

To quantify the clinical impact of the differing sensitivities, each virtual patient in the simulation was classified into a 2-by-2 contingency table based on whether their true SWI count (*Y_SWI_*) and simulated T2* count (*Y_T2_*_∗_) were below or above the five-CMH exclusion threshold (see Table S1). The four cells of this table are defined as follows: cell *a*, *Y_SWI_* ≥5 and *Y_T2_*_∗_ ≥5, representing patients classified as ineligible by both sequences. Cell *b*, *Y_SWI_* ≥5 and *Y_T2_*_∗_ <5, representing patients classified as ineligible by SWI but eligible by simulated T2*-GRE. Cell *c* (reverse discordance): *Y_SWI_* <5 and *Y_T2_*_∗_ ≥5, where patients would be classified as eligible by SWI but ineligible by T2*-GRE. Under this binomial detection model, the simulated T2*-GRE count cannot exceed the SWI-based count. Therefore, cell *c* was retained in the classification framework for completeness but is structurally zero under the primary directional model. Cell *d*, *Y_SWI_* <5 and *Y_T2_*_∗_ <5, representing patients classified as eligible by both sequences.

Based on this classification framework, we calculated two primary outcome measures to assess the misclassification risk from different clinical perspectives: (i) under-detection rate of high CMH burden by T2* (*under-detection rate*), this metric quantifies the proportion of SWI-ineligible patients who would be classified as eligible by simulated T2*-GRE. It is calculated from the row of all SWI-ineligible patients as *b/(a+b)*, corresponding to the conditional probability P(*Y_T2_*_∗_ <5 ∣ *Y_SWI_* ≥5). And (ii) the proportion of discordant cases among T2*-eligible patients (*discordant rate*), which assesses the purity of the patient group cleared for treatment by T2*-GRE. It represents the proportion of patients classified as eligible by T2*-GRE who would have been classified as ineligible by SWI. It is calculated from the column of all T2*-eligible patients as b⁄(b + d), corresponding to the conditional probability P(*Y_SWI_* ≥5 ∣ *Y_T2_*_∗_ <5).

### Probabilistic Sensitivity Analysis

To assess the robustness of our findings and account for the uncertainty in our initial model parameters, we conducted a probabilistic sensitivity analysis (PSA). In a separate probabilistic sensitivity analysis, we additionally allowed *p_detect_* to vary continuously. In 5,000 simulation runs, we simultaneously drew λ and zi from their posterior distributions and *p_detect_*from a uniform distribution spanning 0.3 to 1.0. This approach allowed us to generate 95% credible intervals (CrIs) for our outcome measures, reflecting the combined uncertainty from both the underlying patient CMH distribution and the variability in T2*-GRE sensitivity.

### Exploratory ADNI comparison

As an exploratory external comparison, we analyzed microhemorrhage counts from the Alzheimer’s Disease Neuroimaging Initiative (ADNI) [Mueller2005]. ADNI MRI documentation describes that MRI data include image data, numerical summary data, and tables containing acquisition and quality-control information, and that medical MR findings include cerebral microbleeds and superficial siderosis. We used ADNI *MRIFind* records to identify *MET2starw* series and summarized the number of cerebral microhemorrhage findings for each participant and visit. *MET2starw* was considered a susceptibility-sensitive multi-echo T2*-weighted GRE sequence, but it was not treated as equivalent to SWI [Jack2008;Arani2024].

We selected participants with CDR-GS 0.5 or 1.0, MMSE >=20, amyloid positivity defined by Centiloid >=18 or positive amyloid PET visual read, and available MET2starw-derived microhemorrhage count. When more than one eligible record was available for a participant, the first eligible record was retained.

To compare the ADNI MET2starw-derived distribution with the primary SWI-based distribution, we fitted the same Bayesian right-censored zero-inflated Poisson model. Counts of five or more microhemorrhages were treated as right-censored at the threshold of five. The model estimated lambda, the mean count in the Poisson component, and zi, the zero-inflation probability. This ADNI analysis was considered descriptive because MET2starw is not identical to SWI and because sequence-specific detection sensitivity could not be inferred from this subset.

### Ethics

The study was conducted in accordance with the Declaration of Helsinki, and under ethical approval of the Ethics Committee of the TMIG (R23-117).

## Results

### Patient Characteristics and Model Parameters

The baseline characteristics of the 130 patients whose data were used to build the simulation model are summarized in Table 1. The cohort had a median age of 76 years (IQR, 72–80), and 91 patients (70.0%) were women. The median MMSE score was 25 (IQR, 23–26). Most patients had CDR-GS 0.5 (114/130, 87.7%), followed by CDR-GS 1.0 (15/130, 11.5%). Ninety-four patients (72.3%) eventually initiated anti-amyloid therapy, including 93 who received lecanemab and 1 who received donanemab. A key finding from the SWI imaging data was that a majority of patients, 61.5% (80/130), had zero detectable CMHs. In addition, approximately 7.0% (9/130) of patients had five or more CMHs.

**Table 1.**
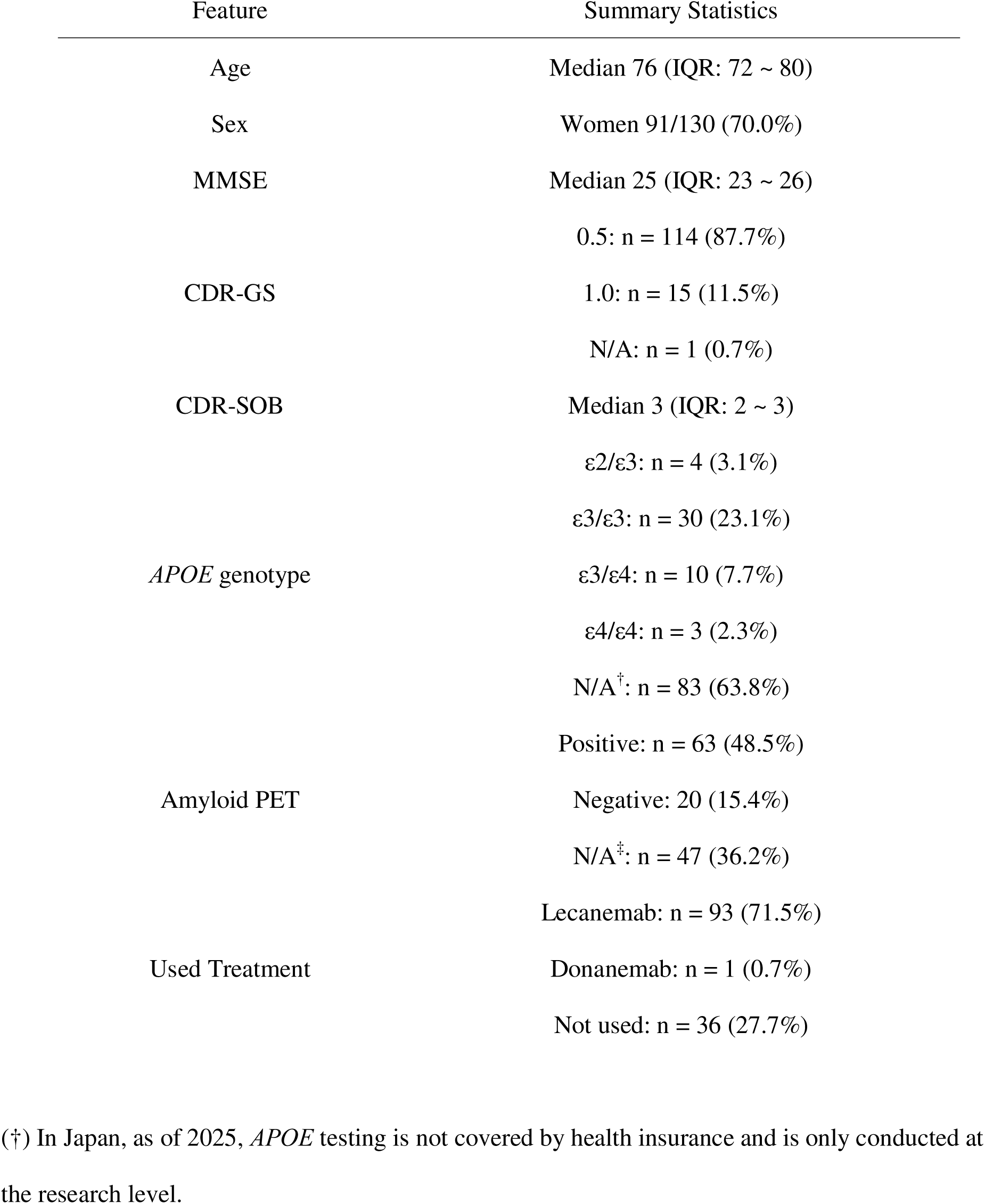
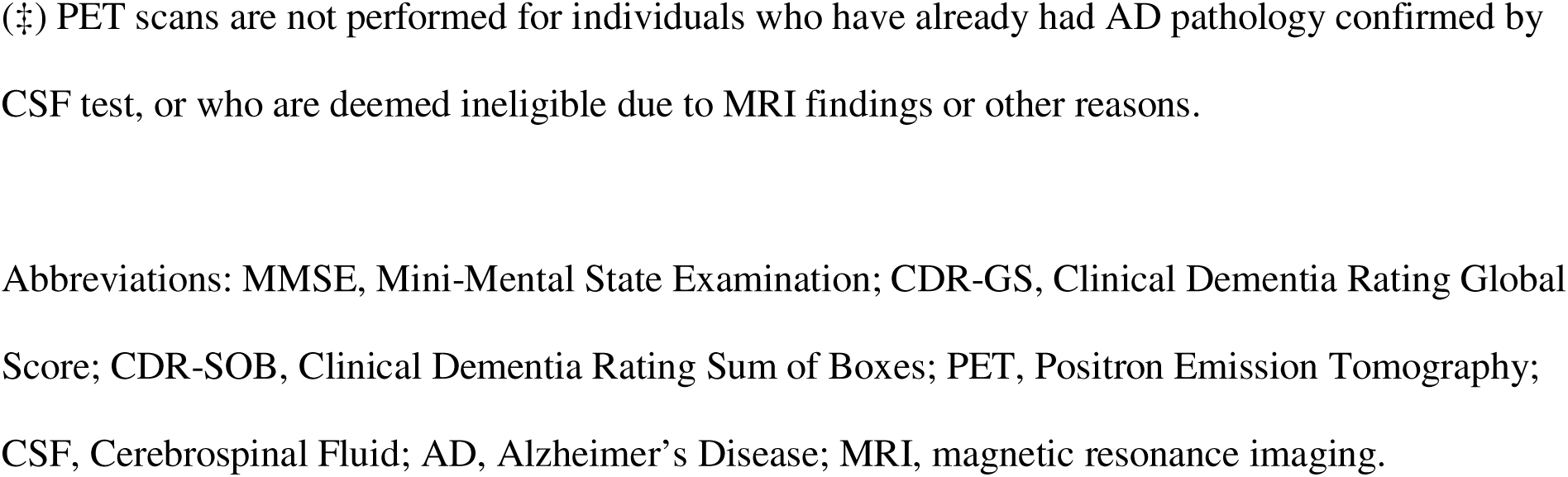
Basic Characteristics of the Analyzed Patients (n = 130)

The Bayesian zero-inflated Poisson model fitted to this data yielded a posterior median for the zero-inflation probability (zi) of 0.57 (95% CrI: 0.47–0.66). The mean CMH count (λ) for the sub-population susceptible to CMHs was estimated to be 2.29 (95% CrI: 1.84–2.81). These parameters, representing the “true” underlying CMH distribution as captured by SWI, formed the basis for our Monte-Carlo simulation. Goodness-of-fit of the obtained model was highly plausible (Figure S1).

### Exploratory ADNI comparison

The exploratory ADNI MET2starw subset included 33 participants (Table S2). The observed microhemorrhage count distribution was sparse and zero-inflated: 21 participants (63.6%) had no microhemorrhages, and 2 participants (6.1%) had five or more microhemorrhages. The observed arithmetic mean count was 0.91. The right-censored zero-inflated Poisson model yielded a posterior median lambda of 2.32 (95% CrI, 1.44-3.40) and zi of 0.588 (95% CrI, 0.384-0.751). These estimates were numerically close to those from the primary SWI-based cohort, in which lambda was 2.29 and zi was 0.57. This exploratory finding suggests that a sparse and zero-inflated susceptibility-sensitive MRI-derived CMH distribution can also be observed in an external research cohort. However, it should not be interpreted as evidence that these parameters are universal across cohorts or sequences. The ADNI subset was small, and MET2starw is a multi-echo T2*-weighted GRE sequence rather than SWI; therefore, this analysis was not used to validate the SWI-derived model or estimate sequence-specific sensitivity.

### Simulation of Misclassification Risk

Our primary simulation results demonstrate that the choice of MRI sequence has a substantial impact on patient eligibility (Figure 2). The (i) under-detection rate of high CMH burden by T2* was notably high (Figure 2A). For instance, at *p_detect_* = 0.60, an illustrative scenario for T2*-GRE compared with SWI, the primary directional model estimated that nearly 80% of patients with five or more CMHs on SWI would be classified as having fewer than five CMHs on T2*-GRE. This estimate should be interpreted as a scenario-based result of the directional binomial detection assumption rather than as an empirical paired-sequence disagreement rate.

**Figure 2.**
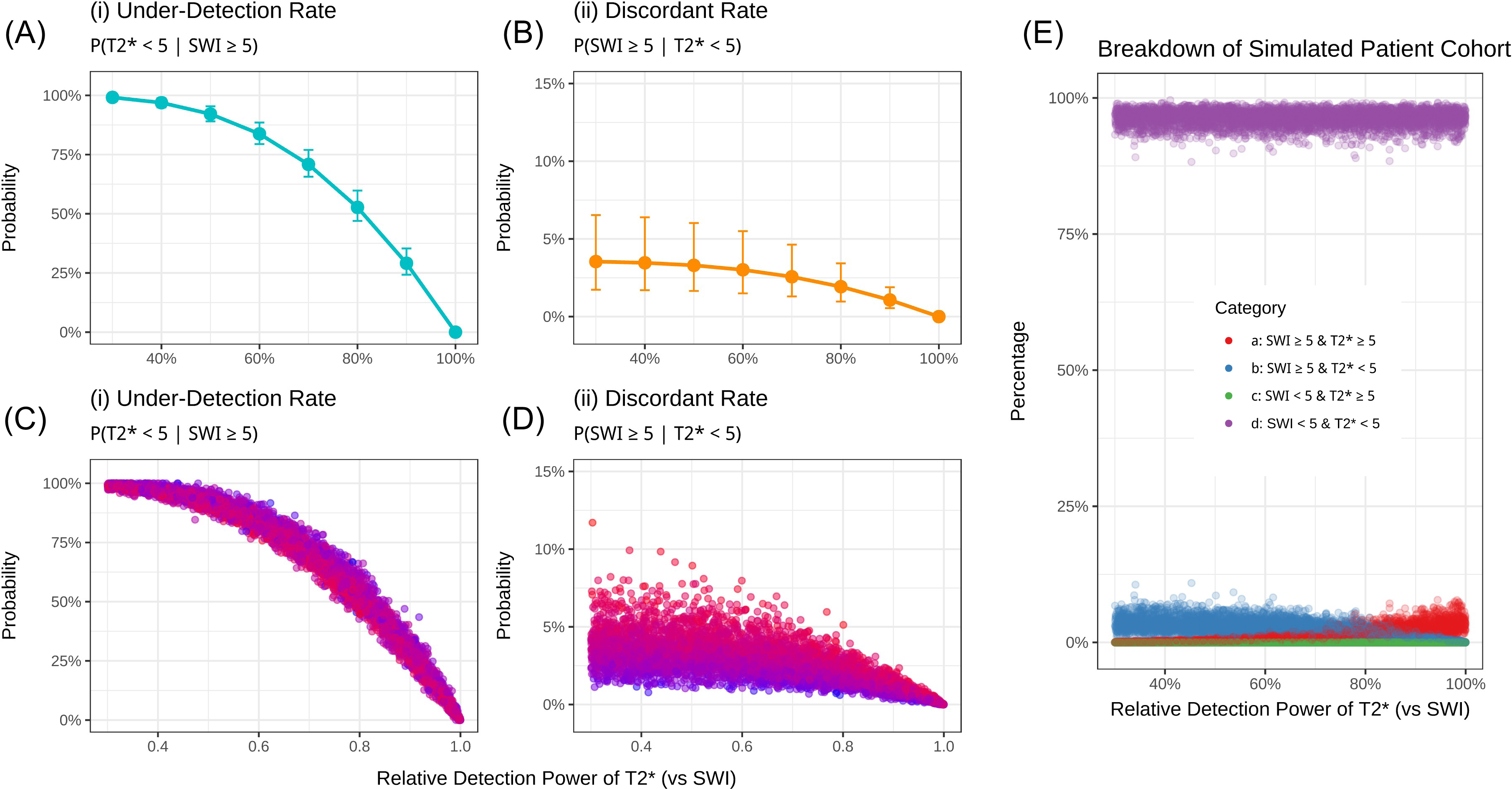
Misclassification Risk Associated with T2*-weighted gradient-recalled echo (T2*-GRE)-Based Screening. The figure illustrates the two primary misclassification probabilities as a function of the relative detection power of T2*-gradient recalled echo (GRE) compared to susceptibility-weighted imaging (SWI). Panels (A) and (B) show the point estimates (medians) and 95% credible intervals derived from the Bayesian posterior distributions. Panels (C), (D), and (E) visualize the full results of the probabilistic sensitivity analysis (PSA), where each point represents one of 5,000 simulations with parameters drawn from their respective distributions. The *Under-Detection Rate* (i: A, C): this metric, calculated as P(*Y_T2*_*< 5 | *Y_SWI_* ≥ 5), quantifies the proportion of SWI-ineligible patients who would be classified as eligible by simulated T2*-GRE. The rate is notably high, decreasing as T2*-GRE sensitivity improves. The *Discordant Rate* (ii: B, D): this metric, calculated as P(*Y_SWI_* ≥ 5 | *Y_T2*_*< 5), represents the proportion of T2*-GRE-eligible patients who would be classified as ineligible by SWI. Breakdown of the simulated patients (E): this panel shows the proportion of the entire virtual cohort falling into each classification category (a, b, c, d) across the range of T2*-GRE sensitivities in the probabilistic sensitivity analysis. Category b (YSWI ≥5 and YT2* <5) represents the main source of sequence-related discrepancy and decreases as T2*-GRE performance improves. Category c is zero under the primary binomial detection model because simulated T2*-GRE counts cannot exceed SWI-based counts. **Abbreviations**: GRE, gradient-recalled echo; SWI, susceptibility-weighted imaging.

The second metric, the proportion of discordant cases among T2*-eligible patients, was smaller but still clinically relevant (Figure 2B). In the illustrative pdetect = 0.60 scenario, approximately 5% of patients classified as eligible by T2*-GRE would have been excluded if SWI had been used.

Because the baseline CMH distribution and relative sequence performance may vary across institutions, we also implemented a static web-based simulator (https://ariah-swi-t2star-simulator.vercel.app/) that allows users to enter local values for zi, λ, and *p_detect_* and obtain classification probabilities based on the same analytical framework. For details, see Figure S2.

### Sensitivity Analysis and Overall Population Impact

The PSA revealed that baseline patient risk influences these misclassification probabilities (Figure 2C, D). In patient populations with a higher underlying CMH burden (i.e., higher λ), the under-detection rate (i) was slightly lower (Figure 2C), while the discordant rate (ii) was higher (Figure 2D). This suggests that in higher-risk cohorts, the pool of patients eligible for T2*-GRE screening is more likely to contain individuals who would be excluded by SWI.

When viewed across the entire patient population (Figure 2E), the vast majority of individuals are correctly classified as eligible (category *d*: *Y_SWI_* <5 and *Y_T2_*_∗_ <5). The proportion of patients in category b (*Y_SWI_* ≥5 and *Y_T2_*_∗_ <5) was the primary source of sequence-related discrepancy, and this group shrinks as T2*-GRE sensitivity improves. Category c (*Y_SWI_ <5* and *Y_T2*_*≥*5*) was zero under the primary binomial detection model, because the model assumed that T2*-GRE could detect only a subset of CMHs visible on SWI.

## Discussion

Our simulation, parameterized with real-world clinical data, demonstrates how the choice of MRI sequence for detecting CMHs can create a clinical dilemma. Under the primary directional detection model, the less sensitive T2*-GRE sequence was associated with a high under-detection rate among patients who would be excluded based on the more sensitive SWI sequence. At the same time, the estimated discordant rate among T2*-GRE-eligible patients was approximately 5%, suggesting that a small but clinically relevant fraction of patients classified as eligible by T2*-GRE would be classified as ineligible by SWI. This discrepancy may affect both safety-oriented screening and equitable access to therapy [Sato2024(PMID:39358780)], creating two legitimate, competing perspectives that clinicians must navigate.

From a patient safety standpoint, using SWI can be viewed as reducing the probability of missed high-burden CMH cases. The adoption of SWI in clinical practice is driven by its superior ability to detect CMHs [Shams2015; Nandigam2009; Cheng2013], providing a more accurate assessment of a patient’s underlying vasculopathic burden. Our model predicts that relying on T2*-GRE could lead to the inclusion of a subgroup of patients with a significant CMH load (≥5 on SWI) that goes undetected. This is reflected in our finding that, under plausible assumptions of T2*-GRE sensitivity, approximately 5% of the treatment-eligible pool would have crossed the exclusion threshold if SWI had been used. The clinical risk of this discordant subgroup remains uncertain, because ARIA risk is influenced not only by CMH count but also by *APOE* genotype, cortical superficial siderosis, white matter disease, amyloid burden, blood pressure, antithrombotic exposure, and treatment regimen [Cogswell2022; Hampel2023]. Therefore, this subgroup should be considered clinically important, but its absolute ARIA risk requires prospective outcome data.

At the same time, an evidence-grounded standpoint highlights that the ≥5 threshold was calibrated on T2*-GRE in the clinical trials; applying the same numeric cutoff to SWI may deny treatment opportunities to patients whose risk would have been acceptable per the trials [Sima2025]. This highlights a core tension in interpretation: a metric can be viewed from a safety standpoint as the ‘probability of missed exclusion by T2*,’ while from a perspective that prioritizes the clinical trial criteria, it suggests a ‘potential loss of treatment opportunity’ from the application of SWI. The Phase 3 trials for lecanemab and donanemab established their safety and efficacy profiles [vanDyck2023; Sims2023], including ARIA incidence rates, using T2*-GRE for screening [Sima2025]. The exclusion criterion of “five or more CMHs” is therefore intrinsically tied to the detection capabilities of T2*-GRE. Consequently, rigidly applying this same numerical threshold to the more sensitive SWI sequence may lead to the exclusion of patients who would have been considered eligible and safely treated within the landmark trials. This raises an implementation concern: applying the same numeric cutoff to a more sensitive sequence may reduce treatment access for patients who would have met trial-based T2*-GRE eligibility criteria. This does not imply that SWI-based screening is inappropriate; rather, it highlights an evidence gap in which improved detection technology and trial-derived numeric thresholds are not directly interchangeable.

This places clinicians and institutions in some difficult position. In Japanese healthcare systems, strict adherence to the OUGs is mandatory for reimbursement [MHLW2023; MHLW2024]. Once an institution adopts SWI as its standard and identifies five or more CMHs, it is obligated to exclude the patient, regardless of what a hypothetical T2*-GRE scan might have shown. This regulatory reality consolidates the loss of treatment opportunity for this discordant group.

Resolving this dilemma requires a further research effort to bridge the evidence gap. The most critical need is for prospective, real-world observational studies that systematically collect baseline imaging data using both SWI and T2*-GRE sequences in patients initiating AATs [Iwatsubo2025]. By correlating these detailed baseline findings with the longitudinal incidence and severity of ARIA, we can begin to quantify the specific risk associated with the discordant group (SWI ≥5, T2* <5). Such studies should also account for lesion phenotype and anatomical distribution, cortical superficial siderosis, white matter disease burden, *APOE* genotype, blood pressure, and antithrombotic exposure. Such data would be foundational for developing evidence-based, sequence-specific and risk-equivalent guidelines. It is possible that the SWI-based threshold corresponding to an equivalent level of risk is higher than five CMHs. Harmonizing these standards is essential for ensuring that safety protocols are both effective and equitable. In the interim, institutions may make an effort to confirm consistency in their imaging protocols to avoid creating local disparities in patient access. For borderline cases, performing both sequences to clarify eligibility could be considered, though the feasibility and cost-effectiveness of this approach are uncertain.

Our work builds upon previous research, such as clinical trial-based estimation [Sima2025], which highlighted that the choice of MRI sequence impacts the distribution of ARIA-H severity. In addition, a recent TRAILBLAZER-ALZ 6 conference presentation [Svaldi2025] reported a head-to-head comparison of SWI and T2*-weighted GRE for ARIA-H detection, providing important paired-sequence context, although these data should be interpreted as conference-level evidence until full publication. Our study advances this literature by using a model fitted to real-world clinical data from a memory clinic to quantify specific, clinically relevant misclassification probabilities—the under-detection rate by T2* and the discordant rate among T2*-eligible—complete with credible intervals to reflect uncertainty.

A notable point is that the discordant rate estimated in our simulation was numerically close to the paired-sequence estimates reported in TRAILBLAZER-ALZ 6 [Svaldi2025], although the definitions were not identical. In that conference slide presentation, 32 of 840 T2*-GRE-eligible participants (3.8%) would have been ineligible by SWI based on microhemorrhage count alone, and 38 of 840 (4.5%) would have been ineligible based on microhemorrhage or cSS criteria. The same presentation reported that clinical dosing recommendations would have changed in 4.9% of participants if decision-making had been based on SWI. These values are consistent with the approximate 5% discordant rate estimated in our simulation. This numerical consistency is notable because our estimate was derived from an independent real-world Japanese screening cohort and a different methodological approach. However, it should be interpreted as triangulating support rather than direct confirmation, because the trial analysis included cSS and dosing recommendations, whereas our model focused only on baseline CMH count.

The exploratory ADNI MET2starw comparison further illustrates why local calibration is important. Although the ADNI subset yielded λ and zi estimates numerically close to those from our SWI-based cohort, this should not be interpreted as evidence that the same parameters apply universally. CMH count distributions may differ by cohort selection, vascular risk profile, scanner field strength, sequence parameters, and reader experience. The web-based simulator was therefore designed to allow institutions to enter their own local parameters rather than directly transferring estimates from our cohort.

Our study has several limitations. First, the model was parameterized using data from a single, specialized academic center in Japan, and its generalizability to other populations, scanner types, or healthcare systems may be limited. The cohort was also selected from patients who had already entered an anti-amyloid therapy screening pathway, and most ultimately initiated treatment. Therefore, the fitted CMH distribution may not represent the broader memory-clinic population or patients screened at earlier stages of referral. Second, our analysis assumes that SWI provides the “true” biological count of CMHs, an assumption that is not always established [Cheng2013]. In addition, the primary binomial model was directional and did not allow T2*-GRE counts to exceed SWI counts; therefore, reverse discordance due to reader variability, artifacts, or lesion mimics was not represented. This simplification should be considered when comparing our estimates with paired-sequence empirical studies. In particular, the high under-detection rate observed in lower-pdetect scenarios should be interpreted as a scenario-based result of the directional binomial model rather than as an empirical estimate of paired-sequence disagreement in routine practice. Third, the relative sensitivity of T2*-GRE is not a fixed value but varies with field strength, vendor, and imaging parameters [Nandigam2009; Cheng2013; Cogswell2022]; our sensitivity analysis explored a range of this variable, but institution-specific values will differ. Although the exploratory ADNI MET2starw analysis showed a similar sparse and zero-inflated distribution, MET2starw is a multi-echo T2*-weighted GRE sequence and not identical to SWI; therefore, this comparison should not be interpreted as an external validation of SWI-derived counts or as an estimate of sequence-specific sensitivity. Finally, our model focused exclusively on the CMH count and did not incorporate other MRI-based exclusion criteria, such as superficial siderosis [Lopes2024], or other ARIA risk modifiers, including *APOE* genotype, white matter disease burden, blood pressure, amyloid burden, antithrombotic exposure, and treatment regimen. Therefore, our estimates should be interpreted as sequence-dependent CMH classification probabilities rather than individualized ARIA risk predictions.

In conclusion, the choice between SWI and T2*-GRE for AAT eligibility screening creates a potential clinical dilemma, pitting the enhanced safety of a more sensitive sequence against the evidence base established in pivotal trials. Our results expose an evidence–technology gap: improved detection (SWI) and trial-defined thresholds (T2*-GRE) point in different operational directions. Until sequence-specific guidance is available, institutions using SWI for eligibility screening should recognize its potential impact on treatment access, apply sequence-specific imaging policies consistently, and document these choices to maintain fairness.

## Acknowledgements

Data collection and sharing for this project was funded by the Alzheimer’s Disease Neuroimaging Initiative (ADNI) (National Institutes of Health Grant U01 AG024904) and DOD ADNI (Department of Defense award number W81XWH-12-2-0012). ADNI is funded by the National Institute on Aging, the National Institute of Biomedical Imaging and Bioengineering, and through generous contributions from the following: AbbVie, Alzheimer’s Association; Alzheimer’s Drug Discovery Foundation; Araclon Biotech; BioClinica, Inc.; Biogen; Bristol-Myers Squibb Company; CereSpir, Inc.; Cogstate; Eisai Inc.; Elan Pharmaceuticals, Inc.; Eli Lilly and Company; EuroImmun; F. Hoffmann-La Roche Ltd and its affiliated company Genentech, Inc.; Fujirebio; GE Healthcare; IXICO Ltd.;Janssen Alzheimer Immunotherapy Research & Development, LLC.; Johnson & Johnson Pharmaceutical Research & Development LLC.; Lumosity; Lundbeck; Merck & Co., Inc.;Meso Scale Diagnostics, LLC.; NeuroRx Research; Neurotrack Technologies; Novartis Pharmaceuticals Corporation; Pfizer Inc.; Piramal Imaging; Servier; Takeda Pharmaceutical Company; and Transition Therapeutics. The Canadian Institutes of Health Research is providing funds to support ADNI clinical sites in Canada. Private sector contributions are facilitated by the Foundation for the National Institutes of Health (www.fnih.org). The grantee organization is the Northern California Institute for Research and Education, and the study is coordinated by the Alzheimer’s Therapeutic Research Institute at the University of Southern California. ADNI data are disseminated by the Laboratory for Neuro Imaging at the University of Southern California.

Some authors’ affiliation (KS, YN, TI), “*Dementia Inclusion and Therapeutics*,” is an endowed department at the University of Tokyo Hospital funded by Effissimo Capital Management Pte Ltd.

## Ethical Considerations

This study was approved by a local ethics committee (TMIG, ID: R23-117).

## Consent to Participate

This study used clinical data obtained under a comprehensive agreement for the use of medical records. Individual informed consent was not required, as the study employed an opt-out approach in accordance with institutional ethical guidelines.

## Consent for Publication

N/A.

## Data Availability

The ADNI data supporting the findings of this study are openly available at (https://ida.loni.usc.edu/). The TMIG data supporting the findings of this study are not publicly available due to privacy or ethical restrictions.

## Declaration of Generative AI and AI-assisted technologies in the writing process

The authors used cloud large language models (ChatGPT and Gemini) for English proofreading. After using these tools, the authors reviewed and edited the content as needed and take full responsibility for the content of the published article.

## Code Availability

The static web-based simulator is available at https://ariah-swi-t2star-simulator.vercel.app/. The source code will be available at https://github.com/kenisatou-tky/ariah-swi-t2star-simulator.

