## Supplementary Materials for "SWI and T2*-GRE Microhemorrhage Counts in Anti-Amyloid Therapy Eligibility: A Real-World-Calibrated Simulation Study"

**Table S1. Agreement/Disagreement Table of the Number of Microhemorrhages between SWI and T2*-GRE**

|  | *Y_T2*_* ≥ 5 | *Y_T2*_* < 5 |
| --- | --- | --- |
| *Y_SWI_* ≥ 5 | *a* | *b* |
| *Y_SWI_* < 5 | *c* | *d* |

Patients were classified according to whether the number of cerebral microhemorrhages was below or above the exclusion threshold of five on SWI and simulated T2*-GRE. Cell a indicates patients classified as ineligible by both sequences, cell b indicates patients with ≥5 microhemorrhages on SWI but <5 on T2*-GRE, cell c indicates patients with <5 on SWI but ≥5 on T2*-GRE, and cell d indicates patients classified as eligible by both sequences. Under the primary binomial detection model, cell c is retained for completeness but is expected to be zero because simulated T2*-GRE counts cannot exceed SWI-based counts.

**Abbreviations**: CMH, cerebral microhemorrhage; GRE, gradient-recalled echo; SWI, susceptibility-weighted imaging.

**Table S2. Exploratory ADNI MET2starw comparison of microhemorrhage count distribution**

|  | Characteristic | Value |
| --- | --- | --- |
| Basic Characteristics | Participants, N | 33 |
|  | Age, years, median (IQR) | 80 (74-82) |
|  | Female sex, n/N (%) | 16/33 (48.5%) |
|  | Education, years, median (IQR) | 16 (14-18) |
|  | Diagnosis: 1_CN, n/N (%) | 2/33 (6.1%) |
|  | Diagnosis: 2_MCI, n/N (%) | 22/33 (66.7%) |
|  | Diagnosis: 3_Dem, n/N (%) | 9/33 (27.3%) |
|  | CDR-GS 0.5, n/N (%) | 29/33 (87.9%) |
|  | CDR-GS 1, n/N (%) | 4/33 (12.1%) |
|  | MMSE, median (IQR) | 27 (24-29) |
|  | Centiloid, median (IQR) | 90.0 (66.0-132.0) |
|  | Positive amyloid PET visual read, n/N (%) | 32/33 (97.0%) |
|  | Met amyloid-positive selection criterion, n/N (%) | 33/33 (100.0%) |
|  | APOE epsilon 4 carrier, n/N (%) among available | 19/32 (59.4%) |
| CMH Characteristics | Observed arithmetic mean CMH count | 0.909 |
|  | Observed zero-count proportion, n/N (%) | 21/33 (63.6%) |
|  | Observed CMH count >=5, n/N (%) | 2/33 (6.1%) |
|  | CMH count 0, n/N (%) | 21/33 (63.6%) |
|  | CMH count 1, n/N (%) | 6/33 (18.2%) |
|  | CMH count 2, n/N (%) | 1/33 (3.0%) |
|  | CMH count 3, n/N (%) | 1/33 (3.0%) |
|  | CMH count 4, n/N (%) | 2/33 (6.1%) |
|  | CMH count >=5, n/N (%) | 2/33 (6.1%) |
|  | Lambda: mean CMH count in the Poisson component, median (95% CrI) | 2.32 (1.44-3.40) |
|  | zi: zero-inflation probability, median (95% CrI) | 0.588 (0.384-0.751) |
|  | Model-implied overall mean count, median (95% CrI) | 0.94 (0.53-1.54) |
|  | Model-implied probability of zero CMH, median (95% CrI) | 63.3% (46.4-77.7%) |
|  | Model-implied probability of CMH count >=5, median (95% CrI) | 3.5% (0.7-10.5%) |

This table summarizes the characteristics, observed MET2starw-derived cerebral microhemorrhage (CMH) count distribution, and Bayesian zero-inflated Poisson model parameters in the exploratory ADNI subset. Participants were selected to approximate the treatment-eligible population by requiring CDR-GS 0.5 or 1.0, MMSE >=20, amyloid positivity defined by Centiloid >=18 or positive amyloid PET visual read, and available MET2starw-derived CMH count in ADNI MRIFind. MET2starw CMH counts of five or more were treated as right-censored at the threshold of five to match the modeling approach used in the primary analysis. Values are shown as median (IQR), n/N (%), or posterior median (95% credible interval), as appropriate. Lambda denotes the mean CMH count in the Poisson component, and zi denotes the zero-inflation probability. MET2starw was not treated as equivalent to SWI; this analysis was intended only as an exploratory external comparison of susceptibility-sensitive MRI-derived CMH count distributions.

**Abbreviations**: ADNI, Alzheimer's Disease Neuroimaging Initiative; CDR-GS, Clinical Dementia Rating Global Score; CMH, cerebral microhemorrhage; CrI, credible interval; MET2starw, multi-echo T2*-weighted imaging; MMSE, Mini-Mental State Examination; SWI, susceptibility-weighted imaging; ZIP, zero-inflated Poisson.

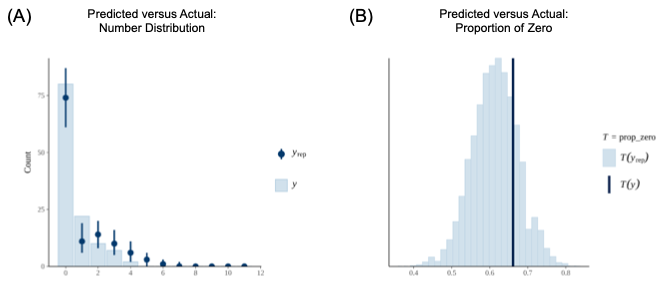
**Figure S1. Goodness-of-Fit of the Model in TMGH cases**

Posterior predictive checks assessing the goodness-of-fit of the Bayesian right-censored zero-inflated Poisson model. (A) The distribution of observed microbleed counts (*y*, light blue bars) is compared to the 95% credible intervals of predicted counts from 1,000 model simulations (*y_rep*, dark blue points and error bars). (B) The observed proportion of zero microbleed counts (T(*y*), dark vertical line) is shown to be highly plausible within the distribution of proportions predicted by the model simulations (T(*y_rep*), histogram).

**Abbreviations**: TMGH, Tokyo Metropolitan Institute for Geriatrics and Gerontology.

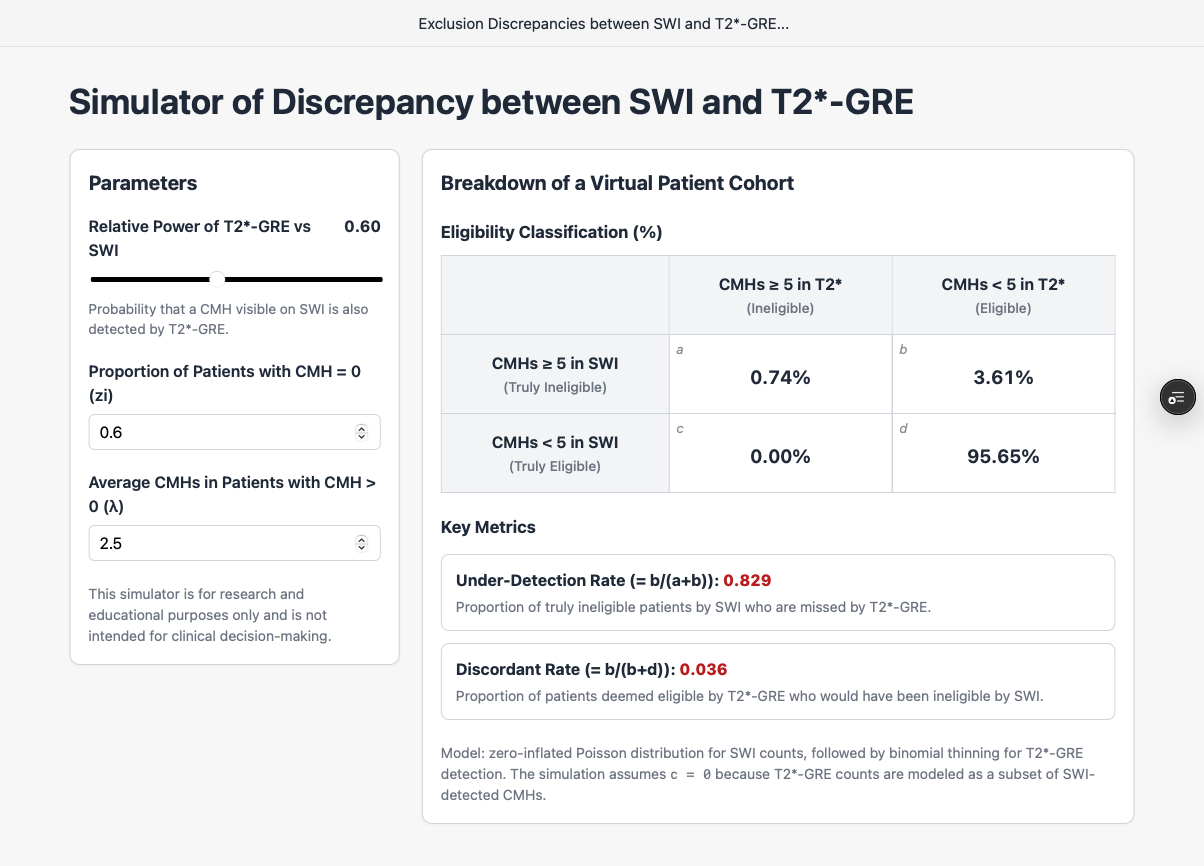
**Figure S2. User Interface of the Web-Based Simulation Tool**

This static web-based simulator (https://ariah-swi-t2star-simulator.vercel.app/) allows users to explore the clinical implications of using T2*-weighted gradient-recalled echo (GRE) for treatment eligibility screening. Users can enter parameters based on their own institution's data:

- Proportion of patients with cerebral microhemorrhage (CMH) = 0 (zi): Enter the proportion of patients with zero microhemorrhages (zi = 0.57 in this study).
- Mean CMH count in the Poisson component (λ): Enter the estimated mean count among patients belonging to the non-structural-zero Poisson component. This parameter is not the observed arithmetic mean among patients with one or more CMHs. In this study, λ was estimated as 2.29.

After setting these parameters, users can adjust the relative detection power of T2*-GRE using the slider to reflect its estimated sensitivity compared with susceptibility-weighted imaging (SWI). The simulator then performs a real-time deterministic calculation to display the percentage of patients falling into each cell of the 2-by-2 contingency table (a, b, c, d). Based on this table, the simulator calculates the two key metrics discussed in this paper: the Under-Detection Rate (= b/(a+b)) and the Discordant Rate (= b/(b+d)). These values are analytical point estimates; they do not account for posterior uncertainty, credible intervals, or the sample size of individual institutions.

**Abbreviations**: GRE, gradient-recalled echo; CMH, cerebral microhemorrhage; SWI, susceptibility-weighted imaging.
